# Endovascular therapy in acute anterior circulation large vessel occlusive patients with low NIHSS (ORIENTAL ENDOLOW): protocol of a multicenter randomized clinical trial

**DOI:** 10.64898/2026.10.03.26364630

**Authors:** Xianjin Shang, Junfeng Xu, Ke Yang, Youqing Xu, Xianjun Huang, Xiangjun Xu, Zi Ye, Jun He, Chong Zheng, Susheng Long, Zhengzhou Yuan, Yufeng Tang, Qian Yang, Jie Xu, Zhiming Zhou

## Abstract

**Introduction:** Mechanical thrombectomy has been well-established as the standard of care treatment for patients with moderate to severe acute ischaemic stroke secondary to large vessel occlusion. However, the clinical value of endovascular treatment (EVT) in patients presenting with minor stroke and low baseline National Institutes of Health Stroke Scale scores remains incompletely defined, with limited high-quality clinical evidence. This protocol outlines the methodology of a randomized trial designed to evaluate whether EVT combined with best medical therapy (BMT) achieves superior functional outcomes relative to BMT alone among patients with minor stroke.

**Methods and analysis:** Eligible patients with acute minor anterior circulation ischaemic stroke and confirmed large vessel occlusion (n = 264) will be randomized via a computer generated block randomization sequence to receive either EVT plus BMT (intervention arm) or BMT alone (control arm).

The primary efficacy outcome is the proportion of participants achieving excellent functional recovery, defined as a modified Rankin Scale score of 0= or 1 at 90 ± 7 days. Primary safety outcomes comprise symptomatic intracranial haemorrhage within 48 hours after randomization (classified according to Heidelberg criteria) and all cause mortality at 90 ± 7 days. Participants will be considered eligible if they satisfy all of the following inclusion criteria: (1) enrolment within 24 hours of stroke symptom onset or time last known well; (2) baseline NIHSS score of 2 - 5 accompanied by at least one focal neurological deficit (including altered consciousness, facial palsy, limb weakness, aphasia or hemispatial neglect); (3) angiographically confirmed acute occlusion of the intracranial internal carotid artery, the M1-segment of the middle cerebral artery, or a dominant M2 branch; (4) a hypoperfusion volume (Tmax > 6 s) ≥ 50 mL on computed tomography or magnetic resonance perfusion imaging. Both intention to treat and per - protocol analyses will be undertaken, and potential sources of heterogeneity in treatment effect magnitude will be explored.

**Ethics and Dissemination:** This study was approved by the ethnic committee from the First Affiliated Hospital of Wannan Medical University (2023-IRB90). The findings of this study will be disseminated through publication in peer-reviewed journals.

**Registration URL:** https://www.clinicaltrial.gov; Unique identifier: NCT06143488.

## INTRODUCTION

Minor ischemic stroke (MIS) is defined as acute ischemic stroke with a National Institutes of Health Stroke Scale (NIHSS) score ≤ 5, accounting for 50–60% of all acute ischemic stroke cases^1–5^. Owing to its subtle clinical manifestations and indolent disease progression, MIS-related risks of early neurological deterioration and long-term recurrent stroke are frequently underestimated in clinical practice^6,7^. Previous studies have reported that early neurological deterioration occurs in 8–30% of MIS, particularly those with concomitant large vessel occlusion (LVO)^2,8,9^, which further raises long-term disability risk^6,10^. Accordingly, MIS is not a benign cerebrovascular event as commonly perceived.

The 2026 AHA/ASA Acute Ischemic Stroke Guidelines provide stratified recommendations for intravenous thrombolysis and dual antiplatelet therapy in MIS. Individualized intravenous thrombolysis is recommended within the 4.5-hour window for patients with disabling deficits including limb dysfunction, aphasia and hemispatial neglect, while evidence for endovascular treatment (EVT) remains insufficient for those with concurrent LVO^11^. Moreover, the 2026 ESO/ESMINT guidelines and international expert consensus recommend against routine EVT for MIS patients with intracranial LVO. EVT is only permitted as a Class IIb recommendation for highly selected patients with disabling deficits or post-thrombolysis neurological worsening, indicating very limited supporting evidence^12,13^.

EVT is the standard therapy for LVO-related acute ischemic stroke, but its approved indications focus primarily on moderate and severe stroke. The role of EVT in minor LVO stroke remains controversial, with no unified clinical consensus established. Several case-control studies demonstrate that EVT achieves rapid recanalization, halts disease progression and improves long-term neurological outcomes^14,15^. However, real-world retrospective studies and meta-analyses yield conflicting results: minor anterior circulation LVO stroke patients undergoing EVT face higher risks of symptomatic intracranial hemorrhage, without improved long-term functional benefits; EVT may even increase odds of procedure-related complications^16,17^. Heterogeneous baseline populations and inconsistent preoperative screening criteria account for these discrepant findings.

Most prior studies enrolled patients solely based on baseline NIHSS scores, ignoring variations in initial symptoms and cerebral perfusion status among individuals. Given expected functional benefits, baseline disabling deficits are presumed core EVT screening criteria for minor LVO stroke. However, prior studies found no efficacy difference between disabling and non-disabling groups^18^. Perfusion imaging has also been applied to optimize reperfusion treatment strategies for this population, with hypoperfusion volume identified as key prognostic factors^15,19,20^. Nevertheless, most relevant studies are single-center retrospective analyses, limited by small sample sizes, non-randomized design and inconsistent imaging assessment standards, leading to low-grade clinical evidence.

Herein, we designed a multicenter, randomized, blinded-endpoint, parallel-controlled clinical trial in Chinese patients with minor LVO stroke. This study aims to compare the efficacy and safety of EVT plus best medical therapy (BMT) versus BMT alone, providing high-quality evidence for clinical decision-making in this patient population.

## METHODS

### Study Design

The Endovascular Therapy versus Best Medical Treatment for Acute Large Vessel Occlusion Stroke with Low NIHSS (ORIENTAL ENDOLOW) study is a multicenter, prospective, open-label, blinded-endpoint, randomized controlled trial.

### Patients population

Patients with acute LVO of intracranial internal carotid artery (ICA) and middle cerebral artery (M1 and main M2 segment), determined by computed tomography angiography (CTA), magnetic resonance angiography (MRA), and/or digital subtraction angiography (DSA), and who meet eligibility criteria and do not meet exclusion criteria will be considered for enrollment at about 16 sites in China. Box 1 lists the inclusion criteria and exclusion criteria.

#### Box 1. Summary of inclusion and exclusion criteria.

##### Inclusion criteria

###### Centre inclusion criteria

1. Equipped with an emergency department and a neurology department for stroke patients.
2. Equipped with an emergency stroke team operating 24 hours a day, 7days a week (24/7).
3. Equipped with commercial perfusion imaging analysis software that can quickly calculate the ischemic hypoperfusion volume.
4. Capable of performing endovascular treatment and intravenous thrombolysis for patients with acute ischemic stroke.

###### Clinical inclusion crtieria

1. Age ≥18 years.
2. Presenting with symptoms underlying acute ischemic stroke.
3. National Institute of Health Stroke Scale (NIHSS) score 2-5 at the time of randomisation, and harboring at least one of the symptom items: (a) impaired consciousness, (b) facial paralysis, (c) limbs weakness, (d) aphasia, (e) neglect.
4. Randomisation can be finished within 24 hours of stroke onset (stroke onset time is defined as last known well time).
5. Informed consent signed.

###### Neuroimaging inclusion criteria

1. CT angiography or MR angiography and/or digital somography angiography proved occlusion of internal carotid artery terminal segment, first [M1] or second main [M2] segment of the middle cerebral artery.
2. Hypoperfusion volume (Tmax >6s) ≥50 mL on CT perfusion or MR perfusion imaging.

##### Exclusion criteria

###### Centre exclusion criteria

1. Centres in which the number of acute ischemic stroke cases treated with endovascular procedures is less than 50 per year.
2. Incapable of complying with the protocol to proceed with the research.

###### Clinical exclusion criteria

1. Patients with modified Rankin Scale scores >1;
2. Patients with artery occlusion caused by other diseases/causes, such as moyamoya disease, vasculitis, radiation-induced vascular disease, fibromuscular dysplasia, and aneurysm or intracranial arteriovenou malformation.
3. Severe comorbid and imbalance conditions, such as heart, lung and renal failure (serum creatinine >246mmol/L(2.8mg/dl)).
4. Having a history of gastrointestinal bleeding, hemorrhagic diseases (such as idiopathic thrombocytopenic purpura) in the past six months.
5. Known bleeding tendency (baseline platelet count <100×10^9^/L/L, etc), or uncontrolled glucose level (defined as persistent baseline glucose level < 50 mg/dL (2.78mmol/L) or > 400 mg/dL (22.2mmol/L)).
6. Patients with contraindications to anesthetics or contrast agents, or a history of drug or alcohol addiction, or mental illness and other conditions that may interfere with preoperative evaluation and lead to poor follow-up compliance.
7. Female who are pregnant, or those of childbearing, potential with positive urine or serum beta Human Chorionic Gonadotropin test.
8. Life expectancy of less than 1 year (such as complicated malignant tumour, serious heart and lung diseases).
9. Participation in other randomised clinical trials that may confound outcome assessment of the trial in the past six months.
10. Other circumstances that the investigator considers inappropriate for participation in the trial or that may pose significant risks to patients (such as inability to understand and/or follow the study procedures).

### Randomisation

The randomization sequence will be generated through a central network randomisation system based on web page platforms or mobile devices. The researcher in each centre will obtain the treatment assignment from the central network randomization system according to the enrollment order, and patients who meet the inclusion and exclusion criteria and provided written informed consent will be randomly assigned (1:1) to EVT+BMT group or BMT group. Central web-based randomization is performed with fixed-size blocks methods to avoid predictable allocation sequences (**Figure 1**).

**Figure 1.**
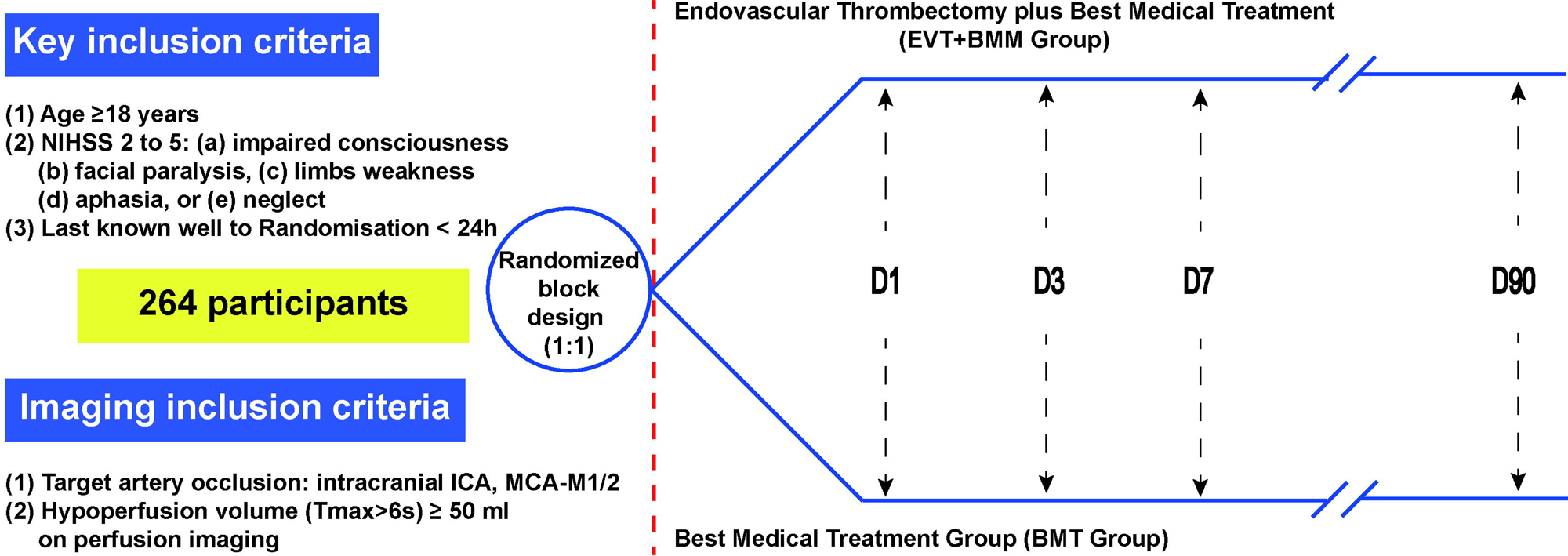
Overview of study design and participant flow.

### Procedural treatment

#### Best Medical Treatment Group (BMT Group)

In accordance with domestic and international guidelines for acute ischemic stroke, patients without indications for intravenous thrombolysis receive early dual antiplatelet therapy with aspirin plus clopidogrel for 21 days, followed by single antiplatelet monotherapy with either agent. Clopidogrel genotyping is performed during treatment; clopidogrel is switched to ticagrelor or cilostazol in poor metabolizers. For patients with severe intracranial arterial stenosis/occlusion or high risk of neurological deterioration, intravenous tirofiban is administered as an optional regimen: a loading dose of 0.4 μg/kg per minute infused over 30 minutes, followed by a maintenance dose of 0.1 μg/kg per minute for 2 to 3 days, with oral antithrombotic agents adjusted thereafter.

Eligible patients with MIS receive priority intravenous thrombolysis with recombinant tissue plasminogen activator (rtPA, 0.9 mg/kg) or tenecteplase (TNK, 0.25 mg/kg). Patients receiving prior thrombolysis are eligible for screening and random enrollment. All thrombolyzed patients follow the same sequential antithrombotic regimens as the participants in the BMT group. A delta NIHSS ≥4 (post-deterioration score minus baseline score) mandates repeat cerebral CT and perfusion imaging. Experienced neurointerventionalists determine the indication for rescue endovascular therapy per clinical guidelines.

#### Endovascular Thrombectomy plus Best Medical Treatment Group (EVT+BMM Group)

EVT for eligible patients with MIS is routinely performed under local anesthesia or conscious sedation, while general anesthesia is applied for elderly, uncooperative or critically ill patients. For patients without prior intravenous thrombolysis, systemic heparinization at 50∼80 U/kg is administered intraoperatively; alternatively, catheters are continuously irrigated with heparinized normal saline to prevent iatrogenic thrombosis.

Endovascular treatment strategies include aspiration thrombectomy, stent retriever thrombectomy, intra-arterial thrombolysis, balloon angioplasty and stent implantation, which may be used in combination as required. For intraoperative intracranial atherosclerotic lesions, balloon angioplasty is performed combined with intra-arterial and intravenous tirofiban. Cerebral reperfusion is monitored for 15 to 30 minutes after recanalization, and rescue stenting is recommended for persistent poor blood flow. After ruling out intracranial hemorrhage on postoperative cranial CT, intravenous tirofiban is continued for 24 to 48 hours. All patients are transferred to the intensive care unit for postoperative vital sign monitoring and follow-up cerebral imaging, with standardized best medical therapy administered throughout the treatment period.

### Follow-up

All participants received serial NIHSS assessments at 24±6 hours, 72±12 hours, day 7±1, or upon hospital discharge. Medication compliance and adverse events were fully documented. Standardized 90-day functional outcome evaluations were performed by two trained independent nurses or physicians blinded to treatment allocation.

### Outcomes

The primary efficacy outcome was excellent functional outcome at 90±7 days after randomisation, as defined by modified Rankin Scale (mRS) score of 0 to 1. Secondary outcomes comprised the following: (1) good functional outcome at 90±7 days as defined by a mRS score of 0 to 2; (2) distribution of mRS scores at 90±7 days; (3) the rate of early neurological deterioration at 24 hours, 72 hours, and 7 days or discharge; (4) the EuroQoL group 5-Dimension 5-Level (EQ-5D-5L) score at 90±7 days.

The primary safety outcome comprised the following: (1) symptomatic intracranial haemorrhage (sICH) within 48 hours (Heidelberg criteria); (2) all-cause mortality at 90±7 days; (3) Any type of ICH within 48 hours (Heidelberg criteria). Secondary safety outcome presented as follows: (1) incidence of all adverse adverse events (including severe adverse events) within 7 days after randomization, including brain herniation, pneumonia, respiratory failure, circulatory failure, stress ulcers, renal failure, acute coronary syndrome, venous thrombosis); (2) procedural and device-related complications (including embolus migration to a new territory, arterial rupture, arterial dissection, stent implantation failure, vascular puncture site complications, vascular spasm, and device malposition).

### Data safety monitoring board

The Data and Safety Monitoring Board (DSMB) conducts regular oversight and review of the entire study enrollment process at predetermined intervals to ensure strict adherence to the study protocol, protect participant safety, and maintain procedural transparency and full ethical compliance. The DSMB consists of academic members independent of study implementation, including professional statisticians. The charter of the DSMB, covering membership qualifications, roles and responsibilities, has been finalized and approved by all members of the DSMB and the Executive Committee. During DSMB meetings, members will formulate formal recommendations, which shall be promptly communicated to the Steering Committee by the DSMB Chair following each meeting.

### Sample size estimates

Sample size calculation for the primary efficacy endpoint was performed based on pooled data derived from the SITS registry and other prospective registry^9,21^, we hypothesized that 50% of patients receiving BMT would attain a excellent 90-day functional outcome, and EVT would raise this rate by 20 percentage points to 70%. With a two-sided alpha level of 0.05, 90% statistical power, and 5% attrition, a total sample size of 264 (132 in each treatment group) were required.

### Statistical Analyses

The efficacy analyses will be performed using the intention-to-treat principle and the missing values will be estimated by multiple imputations. Difference between the two groups in the primary endpoint, proportions of patients with mRS 0-1 at 90 days, will be assessed using the log-binomial regression model (robust Poisson regression, if the model does not converge) with trial centres as a random effect. The risk ratio (RR) and 95% confidence interval will be reported. Predefined subgroup analyses will be performed according to age (<65years vs. ≥65 years), sex (male vs. female), baseline stroke severity (NIHSS 2-3 vs. 4-5), initial clinical symptoms (limb weakness vs. aphasia), stroke etiology (large vessel occlusion vs. cardioembolism vs. others), onset-to-randomization time (<6 hours vs. 6-24hours), occlusion site (intracranial ICA vs. MCA M1 vs. M2), and admission hypoperfusion volume (<70ml vs. ≥70ml). Secondary efficacy outcomes and subgroup analyses were exploratory without multiplicity correction.

An interim analysis is planned when 132 participants (half of the target sample size) have completed the 90-day follow-up with the use of an O’Brien–Fleming spending function to control the overall type I error rate. A P value <0.003 and <0.049 was considered statistically significant in the interim and final analyses, respectively. The trial would be stopped early for efficacy if the primary efficacy boundary was crossed, for futility if the interim results showed that a conclusion about the treatment effect could not be made with even the full sample size, or for safety concerns as judged by the DSMB.

Statistical analyses will be conducted using R (version 4.2+) and SAS 9.4 software.

### Study organization and funding

The study is funded by the Major Scientific Research Project of Anhui Provincial Department of Education (2023AH040256) and the Key Scientific Research Project of Anhui Provincial Health Commission (2024Aa10059). The funding entities were not involved in the study design and will not be involved in the study monitoring, data collection, statistical analyses, interpretation of results, or manuscript writing.

## DISCUSSION

The Oriental ENDOLOW study aims to explore whether EVT improves 90-day excellent functional outcomes compared with BMT in patients with acute anterior circulation LVO and MIS defined by low NIHSS scores. Several preliminary studies have investigated EVT for minor stroke with LVO, additional randomized controlled trials are urgently required to verify whether EVT can mitigate the risks of neurological deterioration and long-term disability in this patient population.

To date, three landmark randomized controlled trials (RCTs) investigating EVT for minor LVO have been launched globally. Among these trials, the MILD-MT Trial (NCT06179017)^22^, — the first completed randomized controlled trial evaluating EVT in patients with MIS and acute anterior circulation large vessel occlusion — recruited 300 Chinese patients with perfusion-confirmed minor anterior circulation LVO and presented its final study data at the 2026 European Stroke Organisation Conference (ESOC). This trial yielded better excellent functional outcomes with EVT, comparable symptomatic intracranial hemorrhage risk across groups, yet a notably higher intracranial hemorrhage rate in the EVT arm. Two ongoing relevant RCTs face notable enrollment delays. The MOSTE trial (France, NCT03796468)^23^, originally scheduled to complete enrollment by 2025, has not published primary outcomes. The ENDOLOW trial(https://clinicaltrials.gov/ct2/show/NCT04167527) remains far from its target enrollment of 1,600 participants, making short-term study completion improbable. Our Oriental ENDOLOW trial initiated patient enrollment in January 2024 and is projected to finish enrollment by December 2027.

This trial limited enrollment to patients with NIHSS scores of 2–5, whereas the other three comparable RCTs recruited minor stroke patients with NIHSS ranging from 0 to 5. This inclusion threshold was determined based on two clinical rationales. First, an NIHSS score ≥ 2 correlates with a higher probability of confirmed LVO in patients with minor stroke^24^. Second, the NIHSS cutoff streamlines bedside patient screening and centralized enrollment across multicenter study sites. Furthermore, we imposed strict eligibility criteria for neurological deficits: enrolled patients must present with at least one of five NIHSS item impairments, including focal limb weakness, aphasia, hemispatial neglect, altered consciousness, and facial palsy. This criterion mirrors current stroke guidelines that prioritize intravenous thrombolysis for disabling minor stroke rather than non-disabling minor stroke, as patients with targeted focal deficits carry elevated long-term disability risks^13^.

For vascular occlusion site inclusion, the trial referenced enrollment protocols from the HERMES collaborative meta-analysis^25^ and aforementioned ENDOLOW trial. Eligible lesions include ICA, MCA M1 segment, and major MCA M2 branch occlusion. Recent ethnicity-specific studies focused on Chinese cohorts have validated the safety and efficacy of EVT for isolated MCA M2 occlusion, further supporting the rationality of including M2 segment lesions^26^. Regarding perfusion imaging criteria, we set a 50 mL threshold for ischemic core hypoperfusion volume. Prior perfusion imaging studies confirmed that hypoperfusion volume exceeding 50 mL is an independent predictor of subsequent neurological deterioration^27^; larger hypoperfusion volumes are also associated with greater clinical benefit from EVT recanalization^20^. It is hypothesized that implementation of the aforementioned more stringent enrollment criteria will facilitate screening and selection of mild cases with the highest probability of achieving favorable therapeutic outcomes. This study has several notable limitations. First, patient recruitment is challenging. Patients with minor stroke often refuse invasive EVT due to mild symptoms, raising risks of randomization cross-over and protocol non-compliance. Second, we excluded patients with NIHSS scores of 0-1 despite confirmed LVO, leaving EVT efficacy for this non-disabling subgroup unclear. Unlike Western populations with predominant cardioembolic minor stroke, East Asian patients mostly suffer from atherosclerotic-related minor stroke. We thus planned prespecified subgroup analyses covering TOAST stroke subtypes (intrinsic atherosclerosis vs. Cardiac embolism), baseline NIHSS stratification (2-3 vs. 4-5), primary neurological deficits (limb weakness vs. aphasia), occlusion sites (ICA vs. MCA M1/2), etc. Third, all participants are recruited from Chinese populations, which restricts the external generalizability of our findings.

## CONCLUSIONS

This randomized controlled trial primarily enrolls patients presenting with disabling minor anterior circulation LVO (NIHSS 2–5). It aims to produce high-quality evidence comparing the functional efficacy and safety of EVT with standalone BMT in this subgroup, in whom the clinical benefit of endovascular therapy has yet to be validated.

## Author Contributions

S**-**X., X-J., Y-K.: Conceptualization, Methodology, Investigation, Data curation, Visualization, Writing - original draft. Z, X-Y., H-X., X-X.: Methodology, Formal analysis, Investigation, Writing - review & editing. H-J, Z-C, L-S., T-Y., Y-Z.: Investigation, Data curation. Z-Z., X-J, Y-Q.: Investigation, Resources, Project administration, Writing - review & editing. All authors read and approved the final version of the manuscript.

## Data availability

Individual participant data that underpin the results from this trial, together with the data dictionary and statistical analysis plan, will be available to qualified researchers on reasonable request after publication of the primary outcome manuscript. Requests will be reviewed by the trial steering committee, and a formal data sharing agreement is required prior to data transfer.

## Supporting information

SPIRIT checklist

## Non-standard Abbreviations and Acronyms

MIS: Minor ischemic stroke
NIHSS: National Institutes of Health Stroke Scale
LVO: large vessel occlusion
EVT: endovascular treatment
ICA: internal carotid artery
sICH: symptomatic intracranial haemorrhage
mRS: modified Rankin Scale

## REFERENCE

1. Johnston SC, Amarenco P, Denison H, et al. Ticagrelor and Aspirin or Aspirin Alone in Acute Ischemic Stroke or TIA. N Engl J Med. 2020;383:207–17.

2. Coutts SB, Ankolekar S, Appireddy R, et al. Tenecteplase versus standard of care for minor ischaemic stroke with proven occlusion (TEMPO-2): a randomised, open label, phase 3 superiority trial. Lancet. 2024;403:2597–605.

3. Xiong Y, Meng X, Jin A, et al. Prourokinase vs Standard Care for Patients With Mild Ischemic Stroke: The PUMICE Randomized Clinical Trial. JAMA Neurol. 2025;82:258–66.

4. Xiong Y, Gu H, Zhao XQ, et al. Clinical Characteristics and In-Hospital Outcomes of Varying Definitions of Minor Stroke: From a Large-Scale Nation-Wide Longitudinal Registry. Stroke. 2021;52:1253–58.

5. Reeves M, Khoury J, Alwell K, et al. Distribution of National Institutes of Health stroke scale in the Cincinnati/Northern Kentucky Stroke Study. Stroke. 2013;44:3211–3.

6. Coutts SB, Modi J, Patel SK, et al. What causes disability after transient ischemic attack and minor stroke?: Results from the CT and MRI in the Triage of TIA and minor Cerebrovascular Events to Identify High Risk Patients (CATCH) Study. Stroke. 2012;43:3018–22.

7. Romano JG, Gardener H, Campo-Bustillo I, et al. Predictors of Outcomes in Patients With Mild Ischemic Stroke Symptoms: MaRISS. Stroke. 2021;52:1995–2004.

8. Heldner MR, Jung S, Zubler C, et al. Outcome of patients with occlusions of the internal carotid artery or the main stem of the middle cerebral artery with NIHSS score of less than 5: comparison between thrombolysed and non-thrombolysed patients. J Neurol Neurosurg Psychiatry. 2015;86:755–60.

9. Mazya MV, Cooray C, Lees KR, et al. Minor stroke due to large artery occlusion. When is intravenous thrombolysis not enough? Results from the SITS International Stroke Thrombolysis Register. Eur Stroke J. 2018;3:29–38.

10. Poisson SN, Nguyen-Huynh MN, Johnston SC, Furie KL, Lev MH, Smith WS. Intracranial large vessel occlusion as a predictor of decline in functional status after transient ischemic attack. Stroke. 2011;42:44–7.

11. Prabhakaran S, Gonzalez NR, Zachrison KS, et al. 2026 Guideline for the Early Management of Patients With Acute Ischemic Stroke: A Guideline From the American Heart Association/American Stroke Association. Stroke. 2026.

12. Turc G, Bhogal P, Fischer U, et al. European Stroke Organisation (ESO) - European Society for Minimally Invasive Neurological Therapy (ESMINT) Guidelines on Mechanical Thrombectomy in Acute Ischaemic StrokeEndorsed by Stroke Alliance for Europe (SAFE). Eur Stroke J. 2019;4:6–12.

13. De Santis F, Foschi M, D’Anna L, et al. Acute Treatment of Disabling and Nondisabling Minor Ischemic Stroke: Expert Guidance for Clinicians. Stroke. 2026;57:549–59.

14. Nagel S, Bouslama M, Krause LU, et al. Mechanical Thrombectomy in Patients With Milder Strokes and Large Vessel Occlusions. Stroke. 2018;49:2391–97.

15. Xue R, Zhong W, Zhou Y, et al. Endovascular Treatment for Minor Acute Ischemic Strokes With Large Vessel Occlusion. J Am Heart Assoc. 2022;11:e027326.

16. Goyal N, Tsivgoulis G, Malhotra K, et al. Medical Management vs Mechanical Thrombectomy for Mild Strokes: An International Multicenter Study and Systematic Review and Meta-analysis. JAMA Neurol. 2020;77:16–24.

17. Safouris A, Palaiodimou L, Nardai S, et al. Medical Management Versus Endovascular Treatment for Large-Vessel Occlusion Anterior Circulation Stroke With Low NIHSS. Stroke. 2023;54:2265–75.

18. Schwarz G, Cascio Rizzo A, Matusevicius M, et al. Reperfusion Treatments in Disabling Versus Nondisabling Mild Stroke due to Anterior Circulation Vessel Occlusion. Stroke. 2023;54:743–50.

19. Shang X, Lin M, Zhang S, et al. Clinical Outcomes of Endovascular Treatment within 24 Hours in Patients with Mild Ischemic Stroke and Perfusion Imaging Selection. AJNR Am J Neuroradiol. 2018;39:1083–87.

20. Seners P, Arquizan C, Fontaine L, et al. Perfusion Imaging and Clinical Outcome in Acute Minor Stroke With Large Vessel Occlusion. Stroke. 2022;53:3429–38.

21. Messer MP, Schonenberger S, Mohlenbruch MA, et al. Minor Stroke Syndromes in Large-Vessel Occlusions: Mechanical Thrombectomy or Thrombolysis Only? AJNR Am J Neuroradiol. 2017;38:1177–79.

22. Wu M, Yi T, Xing P, et al. Endovascular treatment for MILD ischaemic stroke with acute anterior circulation large vessel occlusion: a multicentre, prospective, randomised, clinical trial (MILD-MT) protocol. Stroke Vasc Neurol. 2026;2026 Apr 13:svn-2025-004832.

23. Arquizan C, Lapergue B, Gory B, et al. Evaluation of acute mechanical revascularization in minor stroke (NIHSS score 5) and large vessel occlusion: The MOSTE multicenter, randomized, clinical trial protocol. Int J Stroke. 2023;18:1255–59.

24. Maas MB, Furie KL, Lev MH, et al. National Institutes of Health Stroke Scale score is poorly predictive of proximal occlusion in acute cerebral ischemia. Stroke. 2009;40:2988–93.

25. Goyal M, Menon BK, van Zwam WH, et al. Endovascular thrombectomy after large-vessel ischaemic stroke: a meta-analysis of individual patient data from five randomised trials. Lancet. 2016;387:1723–31.

26. Hu W, Jing X, Chen Z, et al. Endovascular Treatment of Medium-Vessel-Occlusion Strokes. N Engl J Med. 2026;394:1894–904.

27. Gwak DS, Kwon JA, Shim DH, Kim YW, Hwang YH. Perfusion and Diffusion Variables Predict Early Neurological Deterioration in Minor Stroke and Large Vessel Occlusion. J Stroke. 2021;23:61–68.

